# Why Symptoms Linger in Quiescent Crohn’s Disease: Investigating the Impact of Sulfidogenic Microbes and Sulfur Metabolic Pathways

**DOI:** 10.1101/2024.09.08.24313266

**Authors:** Jonathan Golob, Krishna Rao, Jeffrey A. Berinstein, Prashant Singh, William D. Chey, Chung Owyang, Nobuhiko Kamada, Peter D.R. Higgins, Vincent Young, Shrinivas Bishu, Allen A. Lee

## Abstract

**Introduction:** Even in the absence of inflammation, persistent symptoms in patients with Crohn’s disease (CD) are prevalent and worsen quality of life. We previously demonstrated enrichment in sulfidogenic microbes in quiescent Crohn’s disease patients with (**qCD+S**) vs. without persistent GI symptoms (**qCD-S**). Thus, we hypothesized that sulfur metabolic pathways would be enriched in stool while differentially abundant microbes would be associated with important sulfur-metabolic pathways in qCD+S.

**Methods:** We performed a multi-center observational study nested within SPARC IBD. Quiescent inflammation was defined by fecal calprotectin level <150 mcg/g. Persistent symptoms were defined by CD-PRO2. Active CD (**aCD**) and non-IBD diarrhea-predominant irritable bowel syndrome (**IBS-D**) were included as controls.

**Results:** Thirty-nine patients with qCD+S, 274 qCD-S, 21 aCD, and 40 IBS-D underwent paired shotgun metagenomic sequencing and untargeted metabolomic profiling. The fecal metabolome in qCD+S was significantly different relative to qCD-S and IBS-D but not aCD. Patients with qCD+S were enriched in sulfur-containing amino acid pathways, including cysteine and methionine, as well as serine, glycine, and threonine. Glutathione and nicotinate/nicotinamide pathways were also enriched in qCD+S relative to qCD-S, suggestive of mitochondrial dysfunction, a downstream target of H_2_S signaling. Multi-omic integration demonstrated that enriched microbes in qCD+S were associated with important sulfur-metabolic pathways. Bacterial sulfur-metabolic genes, including *CTH*, *isfD*, *sarD*, and *asrC*, were dysregulated in qCD+S. Finally, sulfur metabolites with and without sulfidogenic microbes showed good accuracy in predicting presence of qCD+S.

**Discussion:** Microbial-derived sulfur pathways and downstream mitochondrial function are perturbed in qCD+S, which implicate H_2_S signaling in the pathogenesis of this condition. Future studies will determine whether targeting H_2_S pathways results in improved quality of life in qCD+S.

**Key Messages:** **What is Already Known**

- Even in the absence of inflammation, persistent gastrointestinal symptoms are common in Crohn’s disease.
- The microbiome is altered in quiescent Crohn’s disease patients with persistent symptoms, but the functional significance of these changes is unknown.

**What is New Here**

- Sulfur metabolites and sulfur metabolic pathways were enriched in stool in quiescent Crohn’s disease patients with persistent symptoms.
- Multi-omic integration showed enriched microbes were associated with important sulfur metabolic pathways in quiescent Crohn’s disease patients with persistent symptoms.

**How Can This Study Help Patient Care**

- Strategies to decrease sulfidogenic microbes and associated sulfur metabolic pathways could represent a novel strategy to improve quality of life in quiescent Crohn’s disease with persistent GI symptoms

## Introduction

While modern therapies have improved care in patients with inflammatory bowel disease (IBD), decreasing inflammation does not necessarily improve quality of life (QoL) for many IBD patients. Even in the absence of active inflammation (i.e. ‘quiescent’), up to 46% of IBD patients report persistent symptoms, particularly with Crohn’s disease (CD).^1^ Quiescent CD with persistent gastrointestinal (GI) symptoms (**qCD+S**) results in worse QoL,^2^ higher costs of care,^3^ and increased risk for opioid use.^4^ Furthermore, both the Food and Drug Administration and the European Medicines Agency recommend the use of patient reported outcomes (PROs) as primary endpoints in IBD clinical trials. Thus, understanding drivers of qCD+S is critically important. However, the mechanisms underlying the development of qCD+S are poorly understood, and evidence-based therapies do not currently exist.

Initially, subclinical inflammation was thought to be the primary driver of qCD+S.^5^ However, the prevalence of persistent symptoms is similar in IBD patients with and without deep remission.^6^ Thus, inflammation alone cannot fully explain qCD+S. More recently, our group and others have demonstrated the presence of altered microbial communities in quiescent IBD patients with persistent symptoms.^7–10^ Specifically, we showed significant changes in microbial structure, composition, and potential function in patients with qCD+S compared to quiescent CD without persistent GI symptoms (**qCD-S**) and non-IBD patients with diarrhea-predominant irritable bowel syndrome (**IBS-D**), but not compared with active inflammatory CD (**aCD**). Furthermore, we identified that patients with qCD+S were enriched in sulfidogenic microbes and microbial gene pathways important in sulfur metabolism.^7^ However, the functional consequences of these microbial changes in patients with qCD+S are still unknown.

Dietary sulfur is metabolized by certain members of the intestinal microbiota to produce hydrogen sulfide (H_2_S). Increased H_2_S is known to compromise intestinal barrier function^11^ and potentiate visceral hypersensitivity,^12^ which have been implicated as important pathogenic features in qCD+S.^5,13,14^ Thus, we hypothesized that (1) sulfur metabolic pathways would be enriched in patients with qCD+S compared with qCD-S; and (2) differentially abundant microbes in patients with qCD+S would be associated with sulfur metabolic pathways by integrated multi-omic analysis.

## Materials and Methods

### Patient Cohort

We performed a multi-center observational study nested within the Study of a Prospective Adult Research Cohort with Inflammatory Bowel Disease (SPARC IBD), a large prospective cohort with standardized collection of clinical data, biosamples, and patient reported outcomes (PROs).^15^ Inclusion criteria included an established diagnosis of CD and age ≥18 years old, while exclusion criteria included a history of total colectomy or presence of ileostomy/colostomy. Patients from our prior study^7^ were included for analysis if they had stool samples available for paired whole genome shotgun (WGS) metagenomic sequencing and untargeted metabolomic profiling.

Quiescent disease was defined by fecal calprotectin (FCP) <150 mcg/g, which has a negative predictive value of 86% for excluding endoscopic inflammation in CD^16^ and suggested as a treatment target by consensus guidelines.^17^ Stool samples for FCP, WGS, and metabolomic profiling were collected within 4 weeks of completing CD-PRO2 scores. Persistent symptoms were defined as mean abdominal pain ≥2 and daily liquid stool frequency ≥4 using CD-PRO2, which were suggested as clinically meaningful targets by consensus-based recommendations.^17^

As controls, we included aCD patients from SPARC IBD defined by FCP >150 mcg/g and non-IBD patients meeting Rome IV criteria for IBS-D^18^ recruited at the University of Michigan (UM). All patients provided written informed consent prior to enrollment. The institutional review board at UM gave ethical approval for this work (HUM193179).

### Metagenomic Sequencing and Metabolomic Profiling

WGS metagenomic sequencing was performed as described previously^7^ and analyzed via a combination of MetaPhlAn (v2.8) for compositional analysis and *geneshot* for microbial genomics.^19,20^ Stool samples also underwent global untargeted metabolomic profiling (Metabolon, Morrisville, NC). Briefly, samples underwent ultrahigh performance liquid chromatography-tandem mass spectrometry. Raw data were extracted, peak-identified and QC processed, and quantified using area-under-the-curve (see Supplemental Methods). Zero counts were replaced using a Bayesian imputation method based on multiplicative replacement using the R package {zCompositions} (v1.4.0-1). Metagenomic and metabolomic datasets underwent centered log transformation. Metabolites were annotated by Metabolon’s internal libraries as well as the human metabolome database (HMDB v5.0).^21^

### Statistical Analysis

Continuous variables were compared using a Kruskal-Wallis test followed by Dunn’s test for *post hoc* analyses. A two-tailed *p*<.05 was considered significant while adjustment for multiple comparisons was performed using the Benjamini-Hochberg method (target false discovery rate q<.10). All analyses were performed using R (v4.2.3).

### Association of metabolites with phenotype

Dimensionality reduction of log_2_-transformed metabolites was performed using uniform manifold approximation and projection (UMAP), a non-linear technique shown to outperform other dimensionality reduction techniques for identifying biologically relevant clusters, particularly with multi-omic data, using the R package {umap} (v0.2.10.0).^22,23^ Differences in metabolites between groups were estimated using permutational analysis of variance (PERMANOVA) implemented by the *adonis* function from the R package {vegan} (v2.6-4) using Euclidean distances with 10,000 permutations.

Linear regression was employed to determine log_2_-fold changes in metabolites for patients with qCD+S vs. qCD-S, aCD, and IBS-D. Metabolites were annotated using knowledge-based ‘sub-pathways’ from Metabolon and 2×2 contingency tables were constructed for each sub-pathway. Over-representation analysis was performed using a hypergeometric test. Metabolite set enrichment analysis using the metabolite concentration table was also performed using MetaboAnalyst (v5.0) with pathways annotated using the KEGG pathway database.^24^

### Multi-omic Data Integration

Given the high-dimensionality and multivariate properties of multi-omic datasets, linear regression and/or pairwise correlation for data integration would not be appropriate for data integration. Instead, we employed lasso regression, a machine learning technique, which uses regularization to handle the high-dimensionality of the datasets and perform variable selection using the R package {glmnet} (v4.1-7).^25^ Using microbes that we previously identified as enriched/depleted (absolute log-fold change >2) in qCD+S,^7^ we performed lasso regression to determine specific microbial-metabolite interactions as well as metabolic pathways associated with these microbes. A microbe-wise model was implemented using expression of enriched/depleted microbes as response and abundances of untargeted fecal metabolites as predictors to identify specific metabolites that are associated with enriched/depleted microbes in qCD+S. Leave-one-out-cross-validation was employed to optimize the penalty parameters. Desparsified lasso was used to obtain 95% confidence intervals and *p*-values.^26^ FDR-corrected metabolites underwent over-representation analysis to determine perturbations in metabolic pathways associated with enriched/depleted microbes.

As a complementary and independent analysis, we next performed functional annotation of the WGS shotgun sequence data to gain insight into relevant microbial genes involved in sulfidogenesis in qCD+S. To this end, we investigated the prevalence of sulfur-metabolizing genes in intestinal bacterial genomes in patients with qCD+S vs. qCD-S. Raw reads from WGS shotgun sequencing were assembled into contigs and mapped to reads. To identify sulfidogenic microbial genes, experimentally derived protein coding sequences from the metagenome (via *geneshot*)^20^ were aligned to known genes of sulfidogenic microbial enzymes^27,28^ procured from KEGG via the *diamond* aligner. Gene presence/absence was then compared amongst phenotypes using Fisher’s exact test to evaluate potential contributions of microbial sulfidogenesis to qCD+S.

As an additional complementary technique for multi-omic integration and to identify potential biomarkers for qCD+S, we utilized lasso regression using the R package {tidymodels} (v1.0.0) to determine whether microbial species and/or metabolites could predict patients with qCD+S vs. qCD-S. Data were randomly partitioned with 75% of the data used for model training and 25% held-out for testing. Five-fold cross-validation was employed to estimate model accuracy and to tune hyperparameters. The prediction accuracy of the model was evaluated on the independent test set by calculating the area under the receiver operator characteristic curve (AUC). Variable importance scores were extracted to determine species and/or metabolites most important to the model.

## Results

A total of 374 patients had stool for paired metagenomic sequencing and global metabolomic profiling (**Figure 1A**, **Table 1**). CD-PRO2 scores were in patients with qCD+S were similar to aCD (*p*=.96) but lower in qCD-S (*p*<.00001, **Figure 1B**). Fecal calprotectin (FCP) levels in patients with qCD+S were similar to qCD-S (*p*=.90) but higher in aCD (*p*<.00001, **Figure 1C**). CD-PRO2 scores showed weak correlation with FCP levels in all CD patients (r=.28, **Figure 1D**) and quiescent CD patients only (r=.14, **Figure 1E**).

**Figure 1.**
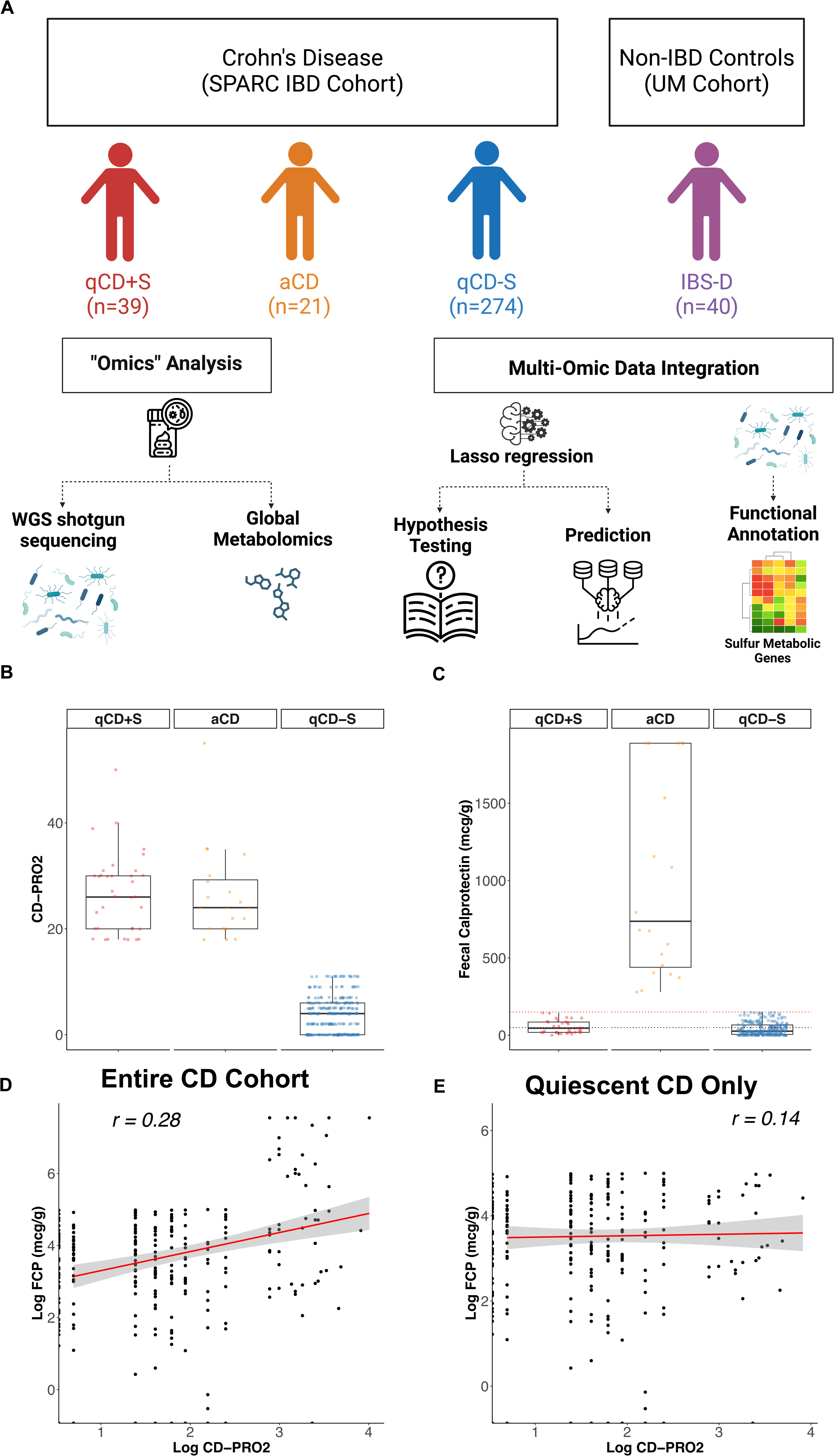
Study design and baseline characteristics. **(A)** Stool samples from 39 patients with qCD+S (*red*), 21 with aCD (*orange*), 274 with qCD-S (*blue*), and 40 with IBS-D (*purple*) underwent paired metagenomic shotgun sequencing and global metabolomic profiling. Multi-omic integration was performed using Lasso penalized regression for hypothesis testing as well as prediction of qCD+S vs. qCD-S. Functional annotation of metagenomic shotgun data was also performed to identify relevant microbial sulfur metabolic genes in qCD+S. (**B**) CD-PRO2 scores in patients with qCD+S (*red*) were similar to aCD (*orange*) but lower in qCD-S (*blue*). (**C**) FCP levels in patients with qCD+S (*red*) were similar to qCD-S (*blue*) but higher in aCD (*orange*). Dotted horizontal *red* and *black* lines indicate cut-off for FCP of 150 mcg/g and 50 mcg/g stool, respectively. There was weak correlation (r=0.28) between log transformed FCP levels and CD-PRO2 scores in (**D**) the entire CD cohort as well as in (**E**) quiescent CD patients only (r=0.14). *Red* line indicates best linear fit with 95% confidence interval (r, Spearman correlation coefficient).

### Fecal Metabolite Characteristics

A total of 1702 fecal metabolites were detected with most metabolites annotated into lipid, amino acid, and xenobiotic super-pathways, as well as dipeptide, arginine and proline, secondary bile acid, and leucine, isoleucine, and valine sub-pathways (**Supplemental Figure 1A, 1C**). The proportion of detected metabolites by super- and sub-pathways are also shown by groups (**Supplemental Figure 1B, 1D)**.

### qCD+S have a distinct fecal metabolome

Examining the relationship between fecal metabolites and phenotype, patients with qCD+S showed a distinct fecal metabolome compared with qCD-S and IBS-D (*p*=.001 for both) but not with aCD (*p*=.07, **Figure 2A**). Variation in stool metabolite profiles showed moderate negative correlation with CD-PRO2 scores in qCD+S (r=-0.30) and aCD (r=-0.34) but only weak negative correlation in qCD-S (r=-0.18, **Figure 2B**). There were 472, 115, and 471 differentially abundant metabolites (q<.10) between patients with qCD+S vs. qCD-S, aCD, and IBS-D, respectively (**Figure 2C, Supplemental Table 1**). Eleven, seven, and fifteen sub-pathways were over-represented (q<.10) in patients with qCD+S vs. qCD-S, aCD, and IBS-D, respectively (**Figure 2D**). Metabolites within cysteine/methionine, bile acid, and fatty acid pathways were amongst the most differentially abundant in qCD+S patients relative to other groups (**Figure 2E**).

**Figure 2.**
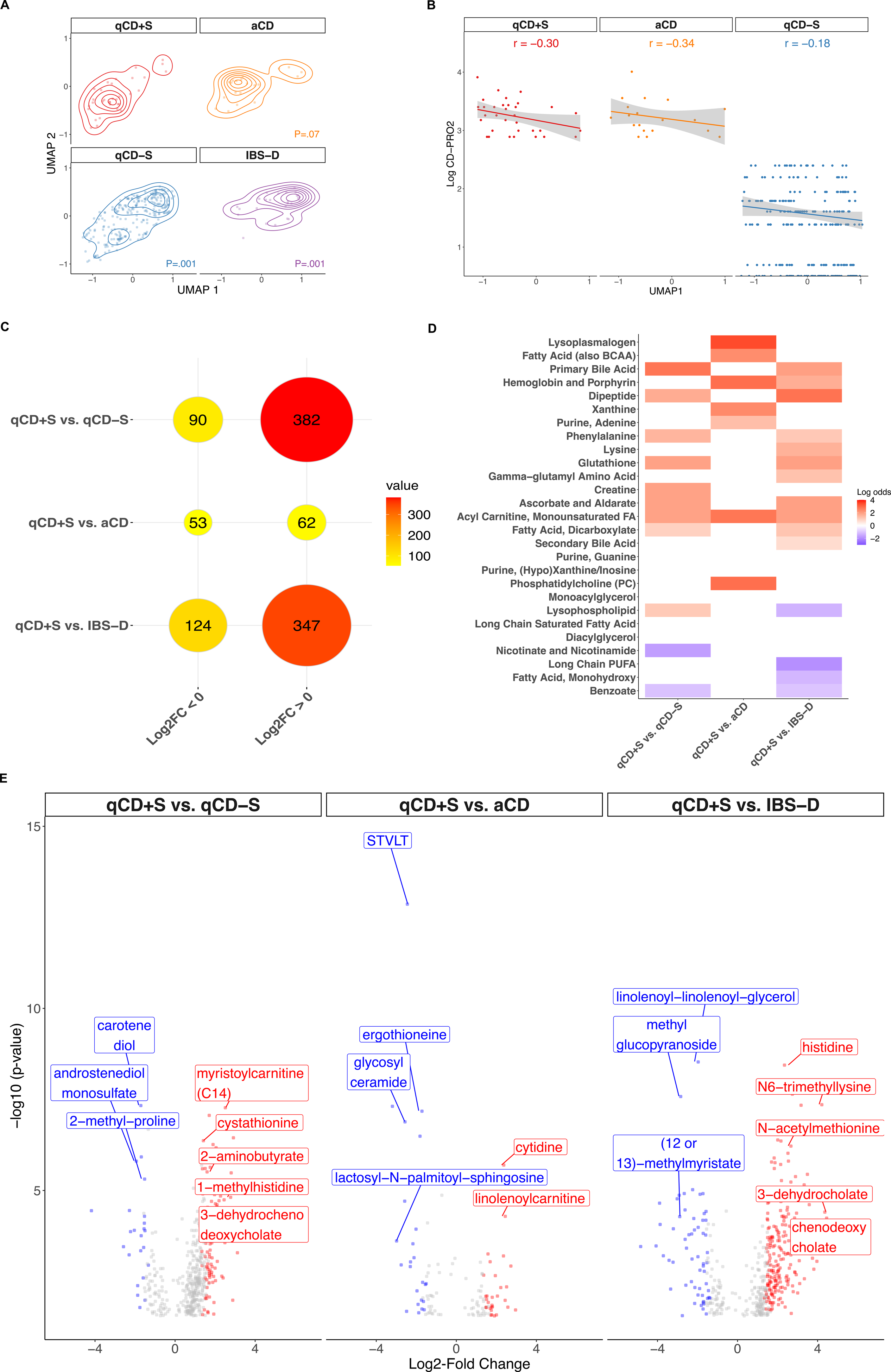
The fecal metabolome in qCD+S is similar to aCD but distinct from qCD-S. (**A**) Uniform manifold approximation and projection (UMAP) overlaid with 2D-contour plot representing ordination of samples according to their metabolite composition. Patients with qCD+S showed significant differences in their fecal metabolite profiles compared to qCD-S and IBS-D by PERMANOVA (*p_adj_*=.001 for both) but not compared with aCD (*p_adj_*=.07). (**B**) There was moderate correlation between log transformed CD-PRO2 scores and the first principal coordinate of UMAP for patients with qCD+S (r=-0.30) and aCD (r=-0.34), but only weak correlation in qCD-S (r=-0.18). *Red* line indicates linear best fit with 95% confidence interval. *r*, Spearman correlation coefficient. (**C**) Balloon plot showing the number of differentially abundant metabolites (q<0.10) by linear regression in qCD+S vs. qCD-S, aCD, and IBS-D. The size and color of each circle are proportional to the number of depleted (log_2_FC <0) or enriched (log_2_FC >0) metabolites in qCD+S vs. other groups with the actual number of metabolites indicated within each circle. (**D**) Heatmap demonstrating sub-pathways that were significantly different (q<.10) in qCD+S relative to other groups. Cell color represents log odds from over-representation analysis, including enriched (*red*), depleted (*blue*), or not significant (*white*) sub-pathways. (**E**) Volcano plot depicting differentially abundant (q<.10) metabolites in qCD+S relative to other groups. *Red*, *blue*, and *grey* dots represent enriched (log_2_FC >1.5 and q<0.10), depleted (log_2_FC <-1.5 and q<0.10), and not significant (abs log_2_FC <1.5 and/or q>0.10) metabolites, respectively. Metabolites within cysteine/methionine (cystathionine, N-acetylmethionine); bile acid (3-dehydrochenodeoxycholate, 3-dehydrocholate, chenodeoxycholate); and fatty acid pathways (myristoylcarnitine, linolenoylcarnitine) were amongst the most differentially abundant metabolites in qCD+S relative to other groups.

### Changes in Fecal Metabolome in Quiescent CD patients with FCP < 50 mcg/g

To determine whether inflammation may have influenced our results, we performed a sensitivity analysis using a more stringent definition for quiescent CD, FCP <50 mcg/g, which has the lowest false-negative rate for inflammation in CD.^29^ The majority of quiescent CD patients (61.5% with qCD+S; 64.6% with qCD-S) had a FCP level <50 mcg/g. Even with this more stringent definition for quiescent CD, we still observed that patients with qCD+S had a distinct fecal metabolome compared with qCD-S (*p*=.001) and IBS-D (*p*=.002) but not aCD (*p*=.07, **Supplemental Figure 2A**). We found 237, 244, and 94 metabolites were differentially abundant in patients with qCD+S vs. qCD-S, aCD, and IBS-D, respectively (**Supplemental Figure 2B**). Many of the most differentially abundant metabolites in qCD+S vs. other groups (**Supplemental Figure 2C**) were similar to our initial analysis (**Figure 2E**).

### qCD+S is enriched in sulfur metabolites

We next investigated our hypothesis that patients with qCD+S are enriched in sulfur metabolites. Of the differentially abundant metabolites, 95, 10, and 95 were sulfur-related metabolites in qCD+S vs. qCD-S, aCD, and IBS-D, respectively (**Figure 3A, Supplemental Table 1**).

**Figure 3.**
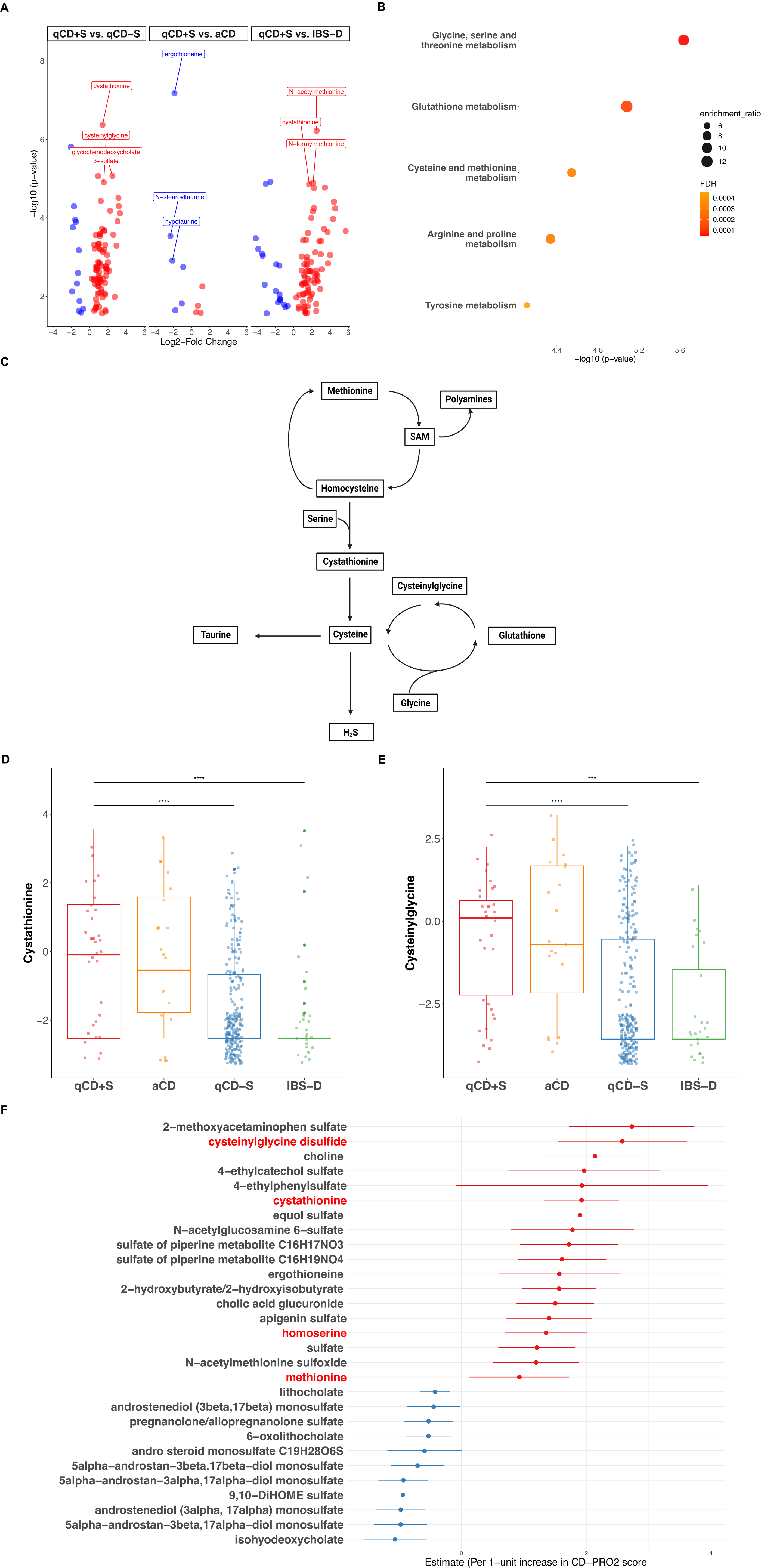
Sulfur metabolic pathways are enriched in qCD+S. (**A**) Volcano plot showing sulfur-related metabolites, including enriched (*red*, log_2_FC >1.5 and q<.10) or depleted (*blue*, log_2_FC <-1.5 and q<.10), in patients with qCD+S relative to other groups. (**B**) Glycine, serine, and threonine; glutathione; and cysteine and methionine were the three most enriched KEGG pathways (q<.10) from metabolite set enrichment analysis of differentially abundant metabolites in qCD+S vs. qCD-S. Color and size of pathway represented by false-discovery rate (FDR) and enrichment ratio (number of observed metabolite hits divided by number of expected metabolite hits within each pathway), respectively. (**C**) Overview of sulfur metabolic pathways in the human gut. Distribution of representative enriched sulfur metabolites, including (**D**) cystathionine and (**E**) cysteinylglycine, in qCD+S relative to other groups. *** *p_adj_* < .0005; **** *p_adj_* < .0001 by Kruskal-Wallis followed by Dunn’s test. (**F**) Association between differentially abundant sulfur-related metabolites (q<.10) and CD-PRO2 score (as a continuous outcome) in qCD+S vs. qCD-S.

We then performed metabolite set enrichment analysis inputting differentially abundant metabolites (q<.10) in qCD+S vs. qCD-S. Glycine, serine and threonine; glutathione; and cysteine and methionine were the three most enriched pathways in qCD+S (**Figure 3B**), which are important sulfur metabolic pathways in the human gut (**Figure 3C**).^27,28^ Cystathionine and cysteinylglycine, both intermediates in cysteine metabolism, were amongst the most enriched metabolites in qCD+S (**Figure 3D-E**). Examining the relationship between CD-PRO2 scores and differentially abundant sulfur-related metabolites (**Figure 3A**), cysteinylglycine disulfide, cystathionine, homoserine, and methionine were among the metabolites most associated with increasing CD-PRO2 scores (indicating worse quality of life) in qCD+S vs. qCD-S (**Figure 3F**).

### Enriched microbes are linked with sulfur metabolism in qCD+S

We next integrated metagenomic and metabolomic datasets to test our hypothesis that enriched microbes in qCD+S were associated with sulfur metabolic pathways. Starting with the most enriched microbes (log-fold change >2) in patients with qCD+S vs. qCD-S^7^ and abundances of untargeted fecal metabolites as predictors (**Figure 4A**), 90 metabolites were significantly associated with enriched microbes (**Supplemental Table 2**). We then performed over-representation analysis and identified taurine and hypotaurine; nicotinate and nicotinamide; cysteine and methionine; and glycine, serine, and threonine as the top metabolic pathways associated with enriched microbes in qCD+S (**Figure 4B**). *Klebsiella oxytoca*, *Bilophila spp.*, and *Prevotella copri* were significantly associated with metabolites in methionine, cysteine, S-adenosylmethionine (SAM), and taurine; primary and secondary bile acids; and nicotinate and nicotinamide while *Streptococcus parasanguinis* was significantly associated with metabolites in glycine, serine, and threonine (**Figure 4C**).

**Figure 4.**
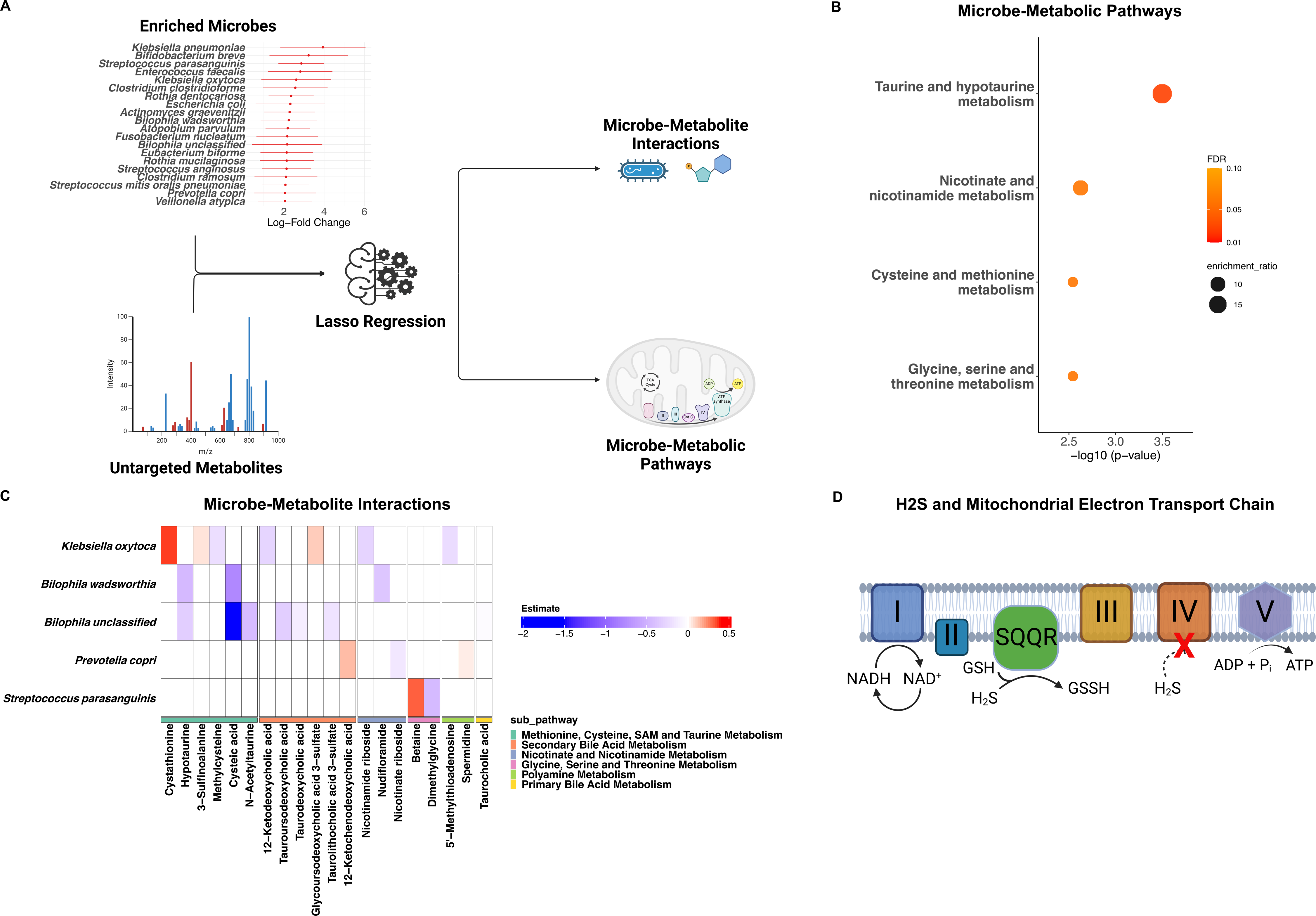
Enriched microbes in qCD+S are associated with sulfur metabolic pathways. (**A**) Schematic of metagenomic-metabolomic data integration using lasso regression. A microbe-wise model was implemented using expression of enriched microbes in patients with qCD+S as response and abundances of untargeted fecal metabolites as predictors. (**B**) Taurine and hypotaurine; nicotinate and nicotinamide; cysteine and methionine; and glycine, serine, and threonine were the top metabolic pathways significantly associated (q<.10) with enriched microbes by over-representation analysis. Color and size of pathway represented by false-discovery rate (FDR) and enrichment ratio (number of observed metabolite hits divided by number of expected metabolite hits within each pathway), respectively. (**C**) Heatmap of significant interactions between enriched microbes and sulfur metabolites. Shading represents estimate (positive, *red*; negative, *blue*) from lasso regression. Metabolites are grouped by sub-pathways (methionine, cysteine, SAM, and taurine, *dark green*; secondary bile acid, *orange*; nicotinate and nicotinamide, *blue*; glycine, serine, and threonine, *pink*; polyamine, *light green*; and primary bile acid, *yellow*). (**D**) The main catabolic pathway of H_2_S in the human gut occurs in mitochondria via the electron transport chain, which is comprised of five protein complexes (designated I-V). Electrons are transferred from NADH to complex I and then further relayed in a series of redox reactions to complexes II, III, and then IV. The electron transport chain generates a proton gradient which is utilized by Complex V to catalyze the formation of ATP from ADP. H_2_S acts as a direct electron donor at the level of sulfide quinone oxidoreductase (SQR), which converts glutathione (GSH) to glutathione persulfide (GSSH). At low concentrations, H_2_S stimulates cellular production of ATP. However, at higher concentrations, H_2_S inhibits complex IV, thereby inhibiting mitochondrial electron transport and results in mitochondrial dysfunction. ADP; adenosine diphosphate; ATP, adenosine triphosphate; H_2_S, hydrogen sulfide; NAD, nicotinamide adenine dinucleotide.

Nicontinate/nicontinamide, precursors for NAD^+^, and glutathione are important co-factors in the mitochondrial electron transport chain, while elevated H_2_S concentrations inhibit mitochondrial function (**Figure 4D**).^30^ Thus, these results suggest interactions between microbial-derived metabolites and host mitochondrial function are of importance in patients with qCD+S.

We similarly examined associations between depleted microbial species (log-fold change <-2) in qCD+S vs. qCD-S and untargeted metabolites. Ninety-three metabolites were significantly associated with these depleted microbes (**Supplemental Table 2**). However, no metabolic pathways were significantly depleted (q>.10).

### Microbial sulfur metabolic genes are dysregulated in qCD+S

To determine relevant microbial pathways for sulfidogenesis in qCD+S, we next annotated the metagenomic shotgun data using known genes of sulfur metabolizing enzymes in the human gut microbiome.^27,28^ We found four genes were significantly differentially abundant in patients with qCD+S vs. qCD-S (q<.10, **Figure 5A-B**).

**Figure 5.**
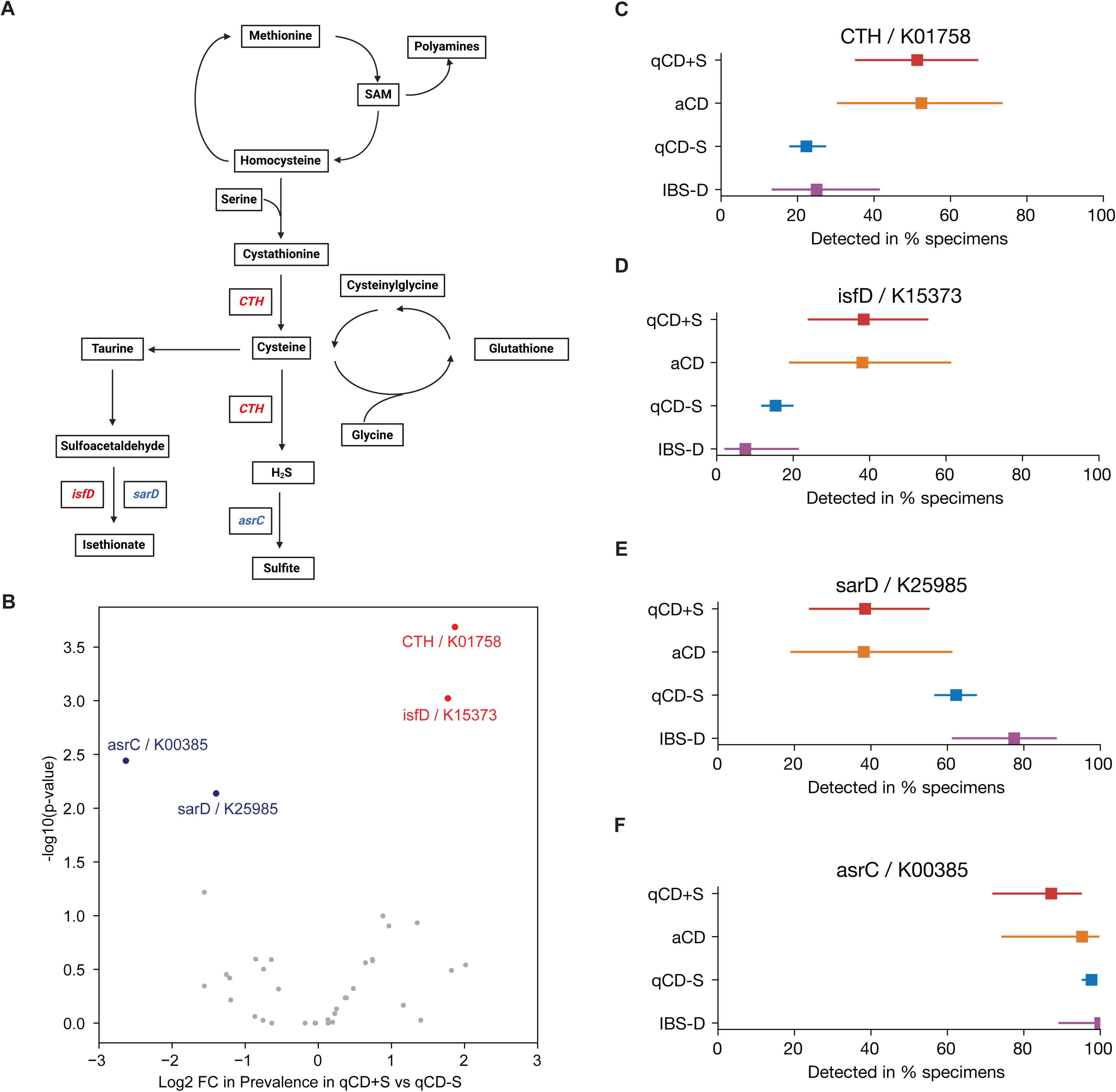
Microbial sulfur metabolic genes are dysregulated in qCD+S. (**A**) Schematic of microbial genes involved in sulfur metabolism that were significantly enriched (*red*) or depleted (*blue*) in patients with qCD+S vs. qCD-S. (**B**) Volcano plot showing significantly enriched (*red*, log_2_FC >1.5 and q<.10), depleted (*blue*, log_2_FC <-1.5 and q<.10), and non-significant (*grey*, absolute log_2_FC <1.5 and/or q>.10) microbial sulfur metabolic genes in qCD+S vs. qCD-S. Distribution of detected microbial genes for (**C**) *CTH*, (**D**) *isfD*, (**E**) *sarD*, and (**F**) *asrC* are demonstrated in qCD+S (*red*), aCD (*orange*), qCD-S (*blue*), and IBS-D (*purple*). Square and bars represent estimate and 95% CI, respectively.

Microbial genes encoding cystathionine gamma-lyase (*CTH*), which converts cystathionine to cysteine and H_2_S, as well as sulfoacetaldehyde reductase (*isfD*), an NAD-dependent enzyme involved in taurine metabolism, were enriched in qCD+S patients compared to qCD-S (**Figure 5C-D**). Anaerobic sulfite reductase subunit C (*asrC*), which catalyzes the conversion of H_2_S to sulfite, and *sarD* (also encoding sulfoacetaldehyde reductase) were significantly decreased in patients with qCD+S vs. qCD-S (**Figure 5E-F**).

### Sulfur metabolites with/without sulfidogenic microbes predict presence of qCD+S

To further test our hypothesis of the importance of sulfidogenic microbes and/or sulfur metabolites as well as to identify potential biomarkers to predict qCD+S, we determined whether sulfidogenic microbes and/or sulfur metabolites could identify patients with qCD+S vs. qCD-S. After randomly splitting the data into train-test set (75%-25%), we trained a lasso regression classifier model using taxonomic composition of enriched microbes (log-fold change >2) as predictors. This model demonstrated modest predictive ability when tested on the held-out test set (AUC 0.66, bootstrapped 95% CI 0.47-0.85) (**Figure 6A**). However, a separate model using sulfur metabolites alone (AUC 0.81, bootstrapped 95% CI 0.69-0.94) showed the best predictive ability (**Figure 6C**) followed by a model combining sulfur metabolites with sulfidogenic microbes (**Figure 6E**, AUC 0.78, bootstrapped 95% CI 0.60-0.96). The top predictors by variable importance plots are shown for each model (**Figures 6B, 6D, 6E**).

**Figure 6.**
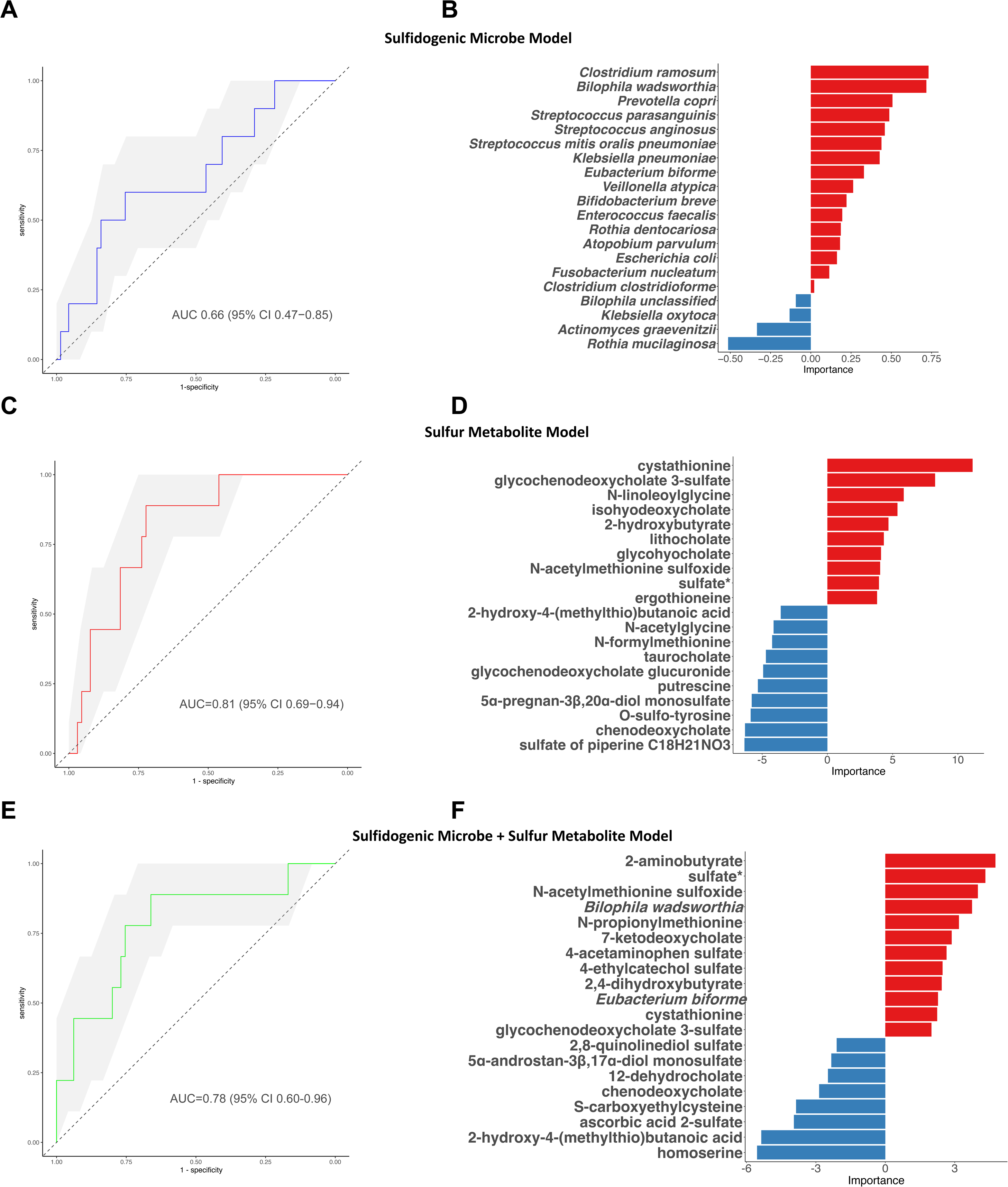
Sulfur metabolites with/without sulfidogenic microbes predict presence of qCD+S. After randomly splitting the data into training (75%) and test (25%) set, a lasso model was trained on (**A**) sulfidogenic microbes (*blue*), (**C**) sulfur metabolites (*red*), and (**E**) combined sulfidogenic microbes-sulfur metabolites (*green*). The top 20 variables by variable importance scores are shown for model with (**B**) sulfidogenic microbes; (**D**) sulfur metabolites; and (**F**) combined sulfidogenic microbes and sulfur metabolites.

## Discussion

In this study combining shotgun metagenomic sequencing and global untargeted metabolomics from a large multi-center cohort, we demonstrated that patients with qCD+S exhibited a distinct fecal metabolome relative to qCD-S and IBS-D but not aCD. Pointing towards potential mechanism, multiple independent but complementary analyses demonstrated enrichment of microbial-derived sulfur metabolic pathways in patients with qCD+S. Glutathione and nicotinate/nicotinamide pathways were also disturbed in qCD+S patients, suggestive of mitochondrial dysfunction, a downstream target of H_2_S signaling. Consistent with our hypothesis, multi-omic integration demonstrated that enriched microbes in qCD+S were associated with sulfur metabolic pathways. Pointing towards relevant microbial pathways, we identified that microbial genes involved in H_2_S, cystathionine, and taurine metabolism were dysregulated in qCD+S patients. Finally, sulfur metabolites with/without sulfidogenic microbes showed good accuracy in identifying patients with qCD+S, suggesting their potential as biomarkers to predict patients at high-risk of developing qCD+S as well as to identify those patients who potentially may respond to sulfur-reducing therapies.

An important observation in our study is that patients with qCD+S showed a distinct fecal metabolome compared with qCD-S but not with aCD. Prior studies have demonstrated that the fecal metabolome largely reflects gut microbial composition and serves as a functional readout of microbial activity.^31^ As such, it is reassuring that our current results mirror our prior findings with fecal metagenomics where patients with qCD+S shared similar microbial diversity, community structure, and membership with aCD but showed significant differences in these indices compared with qCD-S. Collectively, these findings suggest on-going disturbances to the structure and function of the microbiota in qCD+S despite normalization of inflammation.

Several converging lines of evidence in our data point towards dysregulation of sulfur metabolism in qCD+S patients. We demonstrated that important sulfur metabolic pathways, including glycine, serine and threonine; glutathione; cysteine and methionine; and taurine and hypotaurine metabolism, were disrupted in patients with qCD+S. Furthermore, we demonstrated that nicotinate/nicotinamide metabolites, precursors for nicotinamide adenine dinucleotide (NAD), were disturbed in qCD+S. These current findings are consistent with our prior observations that microbial genes encoding cysteine and methionine metabolism, ATP transport, and redox reactions were among the most dysregulated pathways in qCD+S.^7^ Cysteine and methionine are the principal sulfur-containing amino acids in the human gut while glycine and serine are key precursors in the synthesis of methionine and cysteine.^32^ Cysteine can be further metabolized to produce H_2_S, taurine, and/or glutathione. The biological effects of H_2_S are determined by the luminal concentration in the gut while mitochondria are a main target of H_2_S signaling.^33^ At lower concentrations, H_2_S is oxidized by mitochondrial enzymes resulting in ATP synthesis. However, at higher concentrations, H_2_S overwhelms the detoxifying capabilities of the mitochondria, which results in oxidative stress. As glutathione coupled with NAD play critical roles in eliminating reactive oxygen species in mitochondria,^34^ these findings suggest that increased microbial-derived H_2_S and redox-linked mitochondrial dysfunction are key pathogenic features in patients with qCD+S.

Another central finding in our study was the confirmation that enriched microbes in qCD+S were linked to sulfur metabolic pathways. Although we had previously reported that putative sulfidogenic microbes were enriched in patients with qCD+S,^7^ we could not clearly define the functional significance of these microbial changes by metagenomic data alone. By incorporating a multi-omic framework, we identified that enriched microbes in qCD+S were associated with key sulfur pathways, including taurine and hypotaurine; nicotinate and nicotinamide; cysteine and methionine; and glycine, serine, and threonine. We also found that *Klebsiella*, *Prevotella*, and *Bilophila* spp. were central microbial hubs in methionine, cysteine, taurine and nicotinate/nicotinamide metabolism while *Streptococcus* was important in glycine, serine, and threonine pathways. Although H_2_S can be produced endogenously by the host, experiments with germ-free animals have demonstrated that 50-80% of H_2_S production is dependent on the intestinal microbiota.^35^ In particular, *Bilophila* spp. are known to metabolize taurine to H_2_S while cysteine- and methionine-metabolizing bacteria, including *Klebsiella*, *Prevotella*, and *Streptococcus*, are also critical in H_2_S production.^27,28,36^ Increased H_2_S levels have deleterious effects on intestinal barrier function^11^ and visceral nociception,^12^ which are implicated in the pathogenesis of qCD+S.^5,13,14^ Thus, these current analyses strongly support microbial-derived sulfidogenesis as an important pathogenic mechanism in patients with qCD+S.

We further demonstrated that bacterial genes important in H_2_S, cystathionine and taurine metabolism were dysregulated in patients with qCD+S. Cystathionine gamma-lyase (*CTH*), which catalyzes the conversion of cystathionine to cysteine and is the primary source of H_2_S in many microbial species,^37^ was the most enriched microbial gene in qCD+S. Furthermore, *asrC*, which detoxifies H_2_S to sulfite, was significantly depleted in qCD+S. These findings suggest that the microbiota in qCD+S is geared toward producing H_2_S while simultaneously having reduced capacity to catabolize excess H_2_S. Additionally, *isfD* and *sarD*, which both encode sulfoacetaldehyde reductase, were dysregulated in qCD+S. Sulfoacetaldehyde reductase catalyzes the degradation of taurine to isethionate, which can be further metabolized by bacteria, notably *Bilophila*, to produce H_2_S.^38^ Future studies may investigate whether inhibition of specific microbial enzymes, e.g. CTH,^37^ are appropriate targets to reverse sulfidogenesis in patients with qCD+S.

There are several strengths of our work. First, we utilized data from the multicenter, prospective SPARC IBD cohort to improve upon prior small, single-center studies.^8–10^ Second, as race, ethnicity, and geography may influence the structure and function of the microbiome,^39^ utilization of the geographically, racially, and ethnically diverse SPARC IBD cohort allowed us to generalize our findings. Third, patients enrolled in SPARC IBD underwent comprehensive phenotyping, while sample processing and sequencing were highly standardized using a single reference laboratory, which reduces bias and improves reproducibility.^15^ Finally, while our group and others have identified microbial changes in quiescent CD with persistent symptoms, the functional significance of these changes were largely unknown. By incorporating a multi-omic framework, we were able to identify microbial dysregulation of sulfur metabolism as a key pathway in patients with qCD+S.

However, there are limitations to our work as well. First, only a subset of patients had endoscopic evidence of quiescent disease. While prior studies have demonstrated that FCP has good-excellent diagnostic accuracy for endoscopic inflammation in CD,^16^ we cannot exclude the possibility that our results were influenced by occult inflammation.^17,40^ However, our results remained robust even when we restricted our analysis to patients with FCP <50 mcg/g, which has a negative predictive value of 94% for excluding inflammation in CD,^16,29^ and suggests that the changes described herein are largely independent from inflammation. Secondly, while these results support an association between microbial-derived sulfur metabolites and persistent symptoms in quiescent CD, we cannot make any firm conclusions on causation based solely upon these results. Similarly, we do not have longitudinal data to suggest directionality between microbial-metabolite changes and persistent symptoms. Finally, although diet is known to be a major influence on the microbiome and metabolome, we did not have dietary information on CD patients.

In conclusion, we have identified, using multiple converging lines of evidence, that enriched microbes in qCD+S are associated with sulfur metabolic pathways and potential downstream mitochondrial dysfunction. These data may implicate microbial-derived H_2_S in the pathogenesis of intestinal barrier dysfunction and visceral hypersensitivity in qCD+S. However, these results require validation in an independent cohort, particularly with endoscopic confirmation of quiescent inflammation. If confirmed, future studies will determine whether therapies to reverse microbial-derived H_2_S production and/or reduce host mitochondrial dysfunction may improve physiologic abnormalities and improve QoL in qCD+S.

## Data Availability

All data produced in the present study are available upon reasonable request to the IBD Plexus (please see https://www.crohnscolitisfoundation.org/research/plexus/academic-request-for-proposal for full details).

---

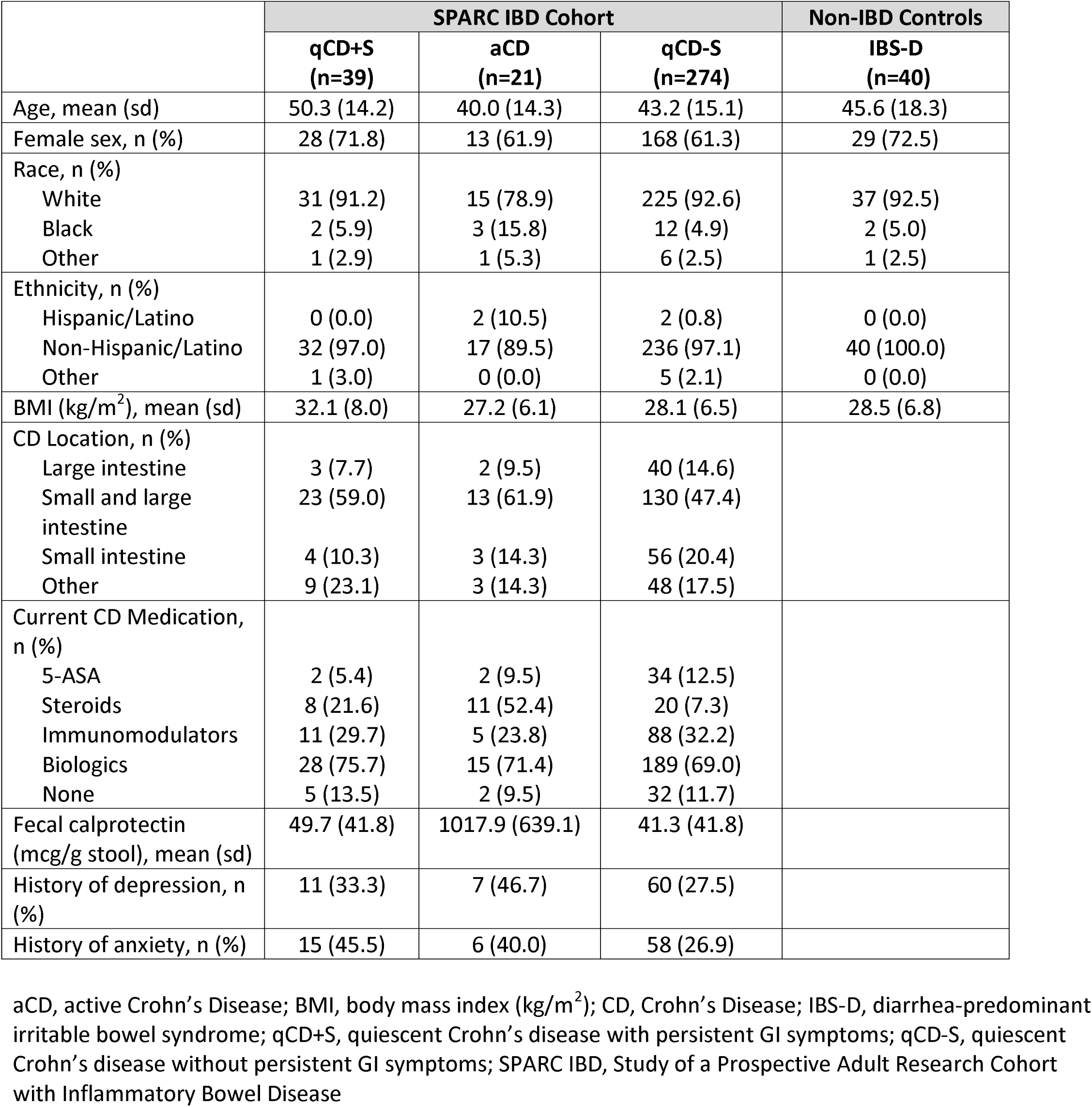

## Notes

**Grant Support:** The results published here are in part based on data obtained from the IBD Plexus program of the Crohn’s & Colitis Foundation. This study was also supported by grants from The Leona M. and Harry B. Helmsley Charitable Trust (to AAL) and the National Institutes of Health grants DK124567 and DK139095 (to AAL); HS027431 (to KR); T32-DK062708, R01-DK125687, R01-DK118154 (to PDHR); AI182787 (to VBY); and DK123403 (to SB).

### Competing Interest Statement

KR is supported in part from an investigator-initiated grant from Merck & Co, Inc.; he has consulted for Seres Therapeutics, Inc., Rebiotix, Inc. and Summit Therapeutics, Inc. VBY has consulted for Vendanta Biosciences, Debiopharm and conducted a speaking engagement sponsored by Aimunne. AAL has consulted for GlaxoSmithKline and Atmo Biosciences. The other authors declare no conflict of interest exist.

### Funding Statement

The results published here are in part based on data obtained from the IBD Plexus program of the Crohns & Colitis Foundation. This study was also supported by grants from The Leona M. and Harry B. Helmsley Charitable Trust (to AAL) and the National Institutes of Health grants DK124567 and DK139095 (to AAL); HS027431 (to KR); T32-DK062708, R01-DK125687, R01-DK118154 (to PDHR); AI182787 (to VBY); and DK123403 (to SB).

### Author Declarations

The institutional review board at the University of Michigan gave ethical approval for this work.

