## Supplementary figures and images for "Why Symptoms Linger in Quiescent Crohn’s Disease: Investigating the Impact of Sulfidogenic Microbes and Sulfur Metabolic Pathways"

### Supplemental Figure 1

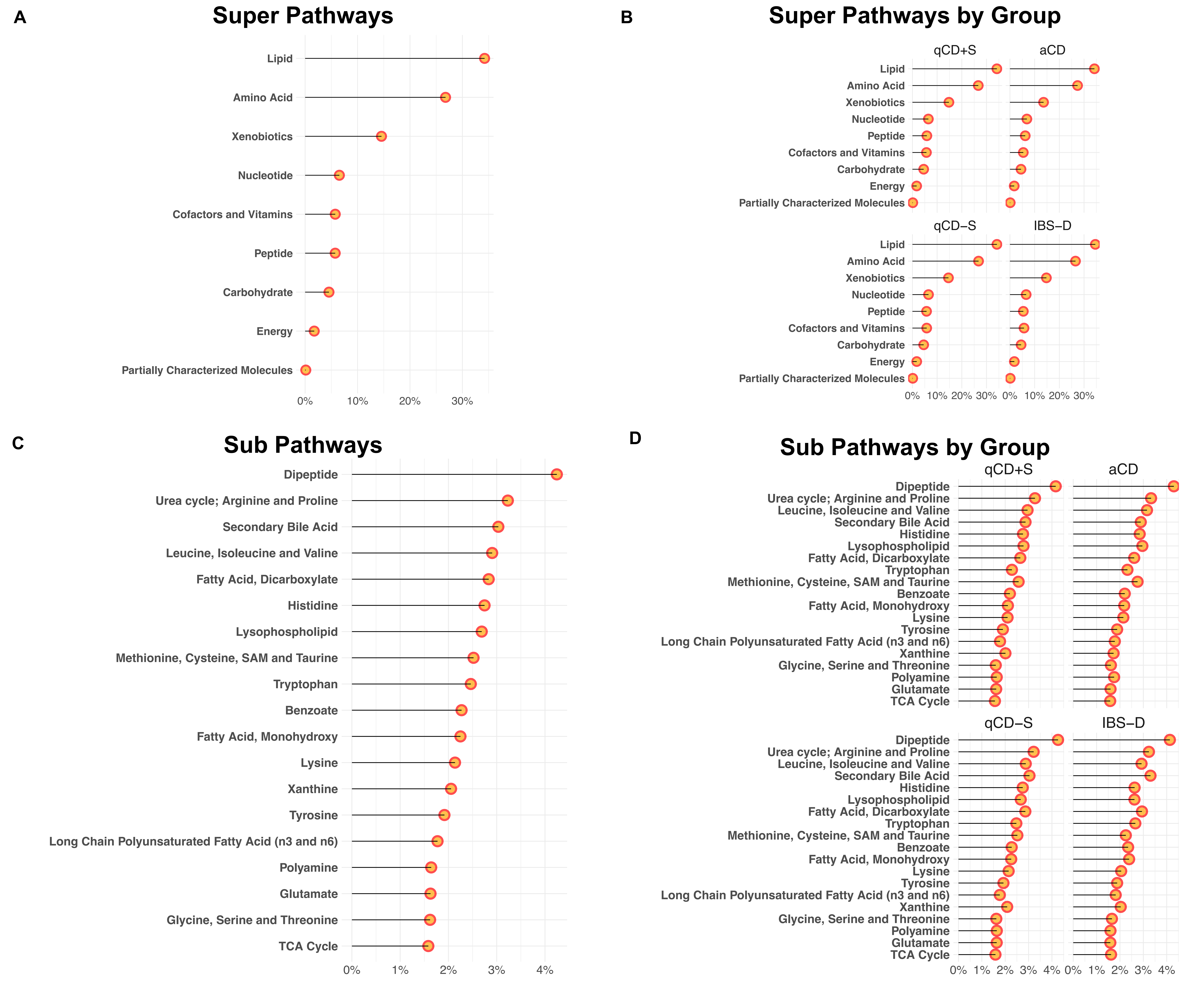

### Supplemental Figure 2

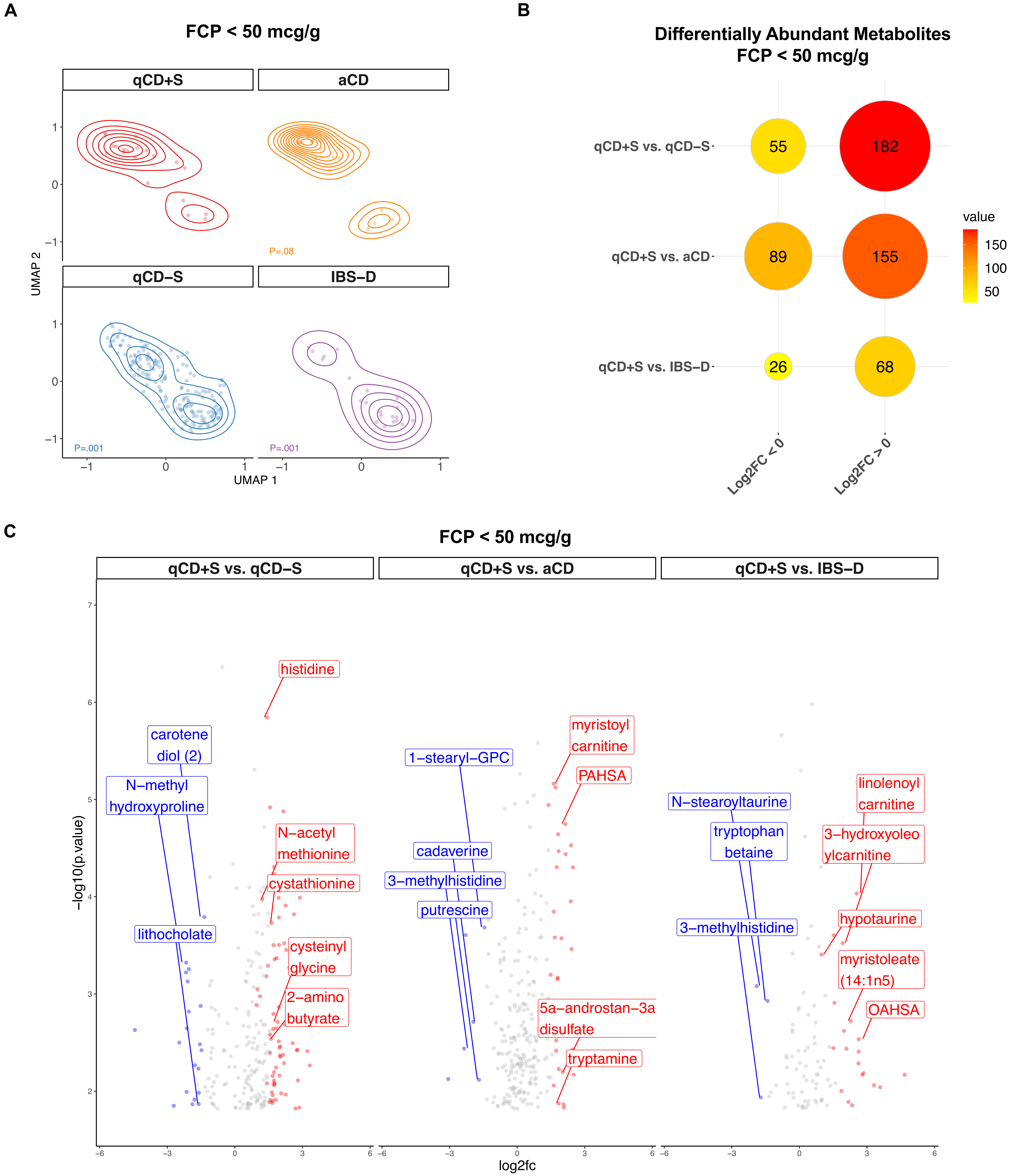
