## Supplemental Material for "Why Symptoms Linger in Quiescent Crohn’s Disease: Investigating the Impact of Sulfidogenic Microbes and Sulfur Metabolic Pathways"

**Supplemental Methods:**

**Untargeted metabolomics platform**

Stool samples collected in 95% ethanol were shipped to Metabolon (Morrisville, North Carolina) for global metabolite profiling. Following receipt, samples were inventoried and immediately stored at -80^o^C. Each sample received was accessioned into the Metabolon LIMS system and was assigned by the LIMS a unique identifier that was associated with the original source identifier only. This identifier was used to track all sample handling, tasks, results, etc. The samples were tracked by the LIMS system. All samples were maintained at -80^o^C until processed.

Samples were prepared using the automated MicroLab STAR® system (Hamilton Company). Several recovery standards were added prior to the first step in the extraction process for QC purposes. Proteins were removed via precipitation with methanol under vigorous shaking for 2 min (Glen Mills GenoGrinder 2000) followed by centrifugation. The resulting extract was divided into five fractions: two for analysis by two separate reverse phase (RP)/UPLC-MS/MS methods with positive ion mode electrospray ionization (ESI), one for analysis by RP/UPLC-MS/MS with negative ion mode ESI, one for analysis by HILIC/UPLC-MS/MS with negative ion mode ESI, and one sample was reserved for backup. Samples were placed briefly on a TurboVap® (Zymark) to remove the organic solvent. The sample extracts were stored overnight under nitrogen before preparation for analysis.

Several types of controls were analyzed in concert with the experimental samples: a pooled matrix sample generated by taking a small volume of each experimental sample served as a technical replicate throughout the data set; extracted water samples served as process blanks; and a cocktail of QC standards that were carefully chosen not to interfere with the measurement of endogenous compounds were spiked into every analyzed sample, allowed instrument performance monitoring and aided chromatographic alignment. Instrument variability was determined by calculating the median relative standard deviation (RSD) for the standards that were added to each sample prior to injection into the mass spectrometers. Overall process variability was determined by calculating the median RSD for all endogenous metabolites (i.e., non-instrument standards) present in 100% of the pooled matrix samples. Experimental samples were randomized across the platform run with QC samples spaced evenly among the injections.

Waters ACQUITY ultra-performance liquid chromatography (UPLC) and a Thermo Scientific Q-Exactive high resolution/accurate mass spectrometer interfaced with a heated electrospray ionization (HESI-II) source and Orbitrap mass analyzer operated at 35,000 mass resolution was employed. One aliquot was analyzed using acidic positive ion conditions. In this method, the extract was gradient eluted from a C18 column (Waters UPLC BEH C18-2.1x100 mm, 1.7 µm) using water and methanol, containing 0.05% perfluoropentanoic acid (PFPA) and 0.1% formic acid (FA). Another aliquot was also analyzed using acidic positive ion conditions but chromatographically optimized for more hydrophobic compounds. In this method, the extract was gradient eluted from the same C18 column using methanol, acetonitrile, water, 0.05% PFPA and 0.01% FA and was operated at an overall higher organic content. Another aliquot was analyzed using basic negative ion optimized conditions using a separate dedicated C18 column. The basic extracts were gradient eluted from the column using methanol and water, however with 6.5mM Ammonium Bicarbonate at pH 8. The fourth aliquot of a given sample was analyzed via negative ionization following elution from a HILIC column (Waters UPLC BEH Amide 2.1x150 mm, 1.7 µm) using a gradient consisting of water and acetonitrile with 10mM Ammonium Formate, pH 10.8. The MS analysis alternated between MS and data-dependent MS^n^ scans using dynamic exclusion with a range covering 70-1000 m/z.

The informatics system consisted of four major components, the Laboratory Information Management System (LIMS), the data extraction and peak-identification software, data processing tools for QC and compound identification, and a collection of information interpretation and visualization tools for use by data analysts. The hardware and software foundations for these informatics components were the LAN backbone, and a database server running Oracle 10.2.0.1 Enterprise Edition.

The purpose of the Metabolon LIMS system was to enable fully auditable laboratory automation through a secure, easy to use, and highly specialized system. The scope of the Metabolon LIMS system encompasses sample accessioning, sample preparation and instrumental analysis and reporting and advanced data analysis. All of the subsequent software systems are grounded in the LIMS data structures. Raw data was extracted, peak-identified and QC processed using Metabolon’s hardware and software. These systems utilize Microsoft’s .NET technologies, which provide active failover and load-balancing. Compounds were identified by comparison to library entries of purified standards or recurrent unknown entities. Metabolon maintains an in-house library based on authenticated standards that contains the retention time/index (RI), mass to charge ratio (m/z), and chromatographic data (including MS/MS spectral data) on all molecules present in the library. Biochemical identifications are based on retention index within a narrow RI window of the proposed identification, accurate mass match to the library +/- 10 ppm, and the MS/MS forward and reverse scores between the experimental data and authentic standards. The MS/MS scores are based on a comparison of the ions present in the experimental spectrum to the ions present in the library spectrum. More than 3300 commercially available purified standard compounds have been acquired and registered into LIMS for analysis on all platforms for determination of their analytical characteristics.

Peaks were quantified using area-under-the-curve. For studies spanning multiple days, a data normalization step was performed to correct variation resulting from instrument inter-day tuning differences. Essentially, each compound was corrected in run-day blocks by registering the medians to equal one (1.00) and normalizing each data point proportionately (termed the “block correction”). For studies that did not require more than one day of analysis, no normalization is necessary, other than for purposes of data visualization.

**Supplemental Figure Legend:**

**Supplemental Figure 1. The number of annotated fecal metabolites is similar in qCD+S compared to other groups.** The number (percent) of annotated metabolites is shown grouped by super pathways using Metabolon’s pathway knowledge base for the (**A**) entire cohort and (**B**) by groups. The top 20 sub-pathways are also shown for the (**C**) entire cohort and (**D**) by groups.

**Supplemental Figure 2. Patients with qCD+S exhibit a distinct fecal metabolome, even when using a more stringent definition for quiescent CD.** (**A**) Quiescent CD patients were restricted to those with FCP <50 mcg/g, while aCD and IBS-D were included as before. The fecal metabolome in patients with qCD+S was distinct from qCD-S and IBS-D (*P_adj_*=.001 for both) but not compared with aCD (*P_adj_*=.08). (**B**) Balloon plot showing the number of differentially abundant metabolites (q<0.10) in patients with qCD+S vs. qCD-S, aCD, and IBS-D. Size and color of each circle are proportional to the number of depleted (log_2_FC < 0) or enriched (log_2_FC > 0) metabolites in qCD+S vs. other groups with the actual number of metabolites indicated within each circle. (**C**) Metabolites within cysteine/methionine (cystathionine, N-acetyl methionine, cysteinyl glycine) and taurine (hypotaurine, N-stearoyltaurine) pathways were amongst the most differentially abundant metabolites in patients with qCD+S vs. other groups.
